# Pharmacological weight loss with incretin-based therapies does not result in a disproportionate loss of muscle mass or function in obese mice and humans

**DOI:** 10.1101/2025.07.28.25332295

**Authors:** Henning Tim Langer, Natalie K. Gilmore, Chris M. T. Hayden, Julien Roux, Bruno Bariohay, Thaïs Rhouquet, Manar Awada, Julie Marcotorchino, Lorrine Bournot, Elizabeth Nunn, Paul M. Titchenell, Daniela Liskiewicz, Timo D. Müller, Oluwaseun Anyiam, Philip J. Atherton, Iskandar Idris, Natalia Haritonow, Kristina Norman, Ursula Müller-Werdan, Keith Baar

## Abstract

The new generation of incretin-based therapies are potent anti-obesity medications (AOMs) that offer the first non-surgical treatment for 936 million patients globally suffering from being overweight or obese[1]. However, clinical data suggest that incretin-mimetics could cause a disproportionate decrease in lean body mass (LBM) [2, 3], raising a concern for deterioration of skeletal muscle and acceleration of sarcopenic obesity[4]. Unfortunately, muscle mass and function are not routinely assessed in obesity studies and original data on the matter remains sparse. In this work, we conducted various pre-clinical studies and a proof-of-concept clinical trial to examine how skeletal muscle is affected by AOMs. We found that in mice with diet-induced obesity (DIO), incretin-based therapies result predominantly in a substantial decrease in fat mass alongside a small but significant decrease in LBM. Among the lean tissues, the decrease in liver mass exceeded the change in muscle mass robustly. While absolute muscle mass did decrease, relative muscle mass (i.e., the muscle mass to body weight (BW) ratio) improved significantly. Similarly, we found that absolute muscle strength decreased mildly but increased relative to the BW of mice. The relative preservation of muscle was also associated with marked improvement in running performance. Additionally, during a scenario of extreme muscle wasting (i.e., immobilization), DIO mice on incretin-based therapies did not experience more muscle loss than calorie-matched, pair-fed mice. Finally, in our clinical proof-of-concept trial, patients on AOMs significantly decreased BW, which was accompanied by a mild decrease in absolute LBM but an improvement in relative LBM. Muscle function as indicated by maximum voluntary contraction (MVC) did not decrease. Overall, these data suggest that in middle-aged obese mice and men, incretin-based therapies do cause a mild decrease in absolute muscle mass and strength that is offset by a more pronounced decrease in fat and liver mass, resulting in an improved muscle to BW ratio, function, and mobility.

## Main

Incretin-based drugs are powerful treatments for obesity. Over the course of 68 to 72 weeks, once weekly injections of semaglutide, a glucagon-like peptide-1 receptor agonist (GLP-1RA), and tirzepatide, a dual agonist for GLP-1 and glucose-dependent insulinotropic polypeptide (GIP), resulted in 12 to 19% reductions in BW compared to placebo, respectively [2, 3]. However, both studies found that a significant portion of the weight loss was achieved through reductions in LBM. For example, roughly 40% of the weight loss in the Wilding study stemmed from LBM[2]. In contrast, LBM is commonly thought to contribute only ∼25% during physiological weight loss, coined the “quarter fat-free mass (FFM) rule”[5]. As such, the concern of a disproportionate loss of LBM, muscle mass and muscle function with incretin-based drugs has spurred tremendous investments by the pharmaceutical industry[6] and the publication of review articles [4, 7]. Despite this great interest in the matter, primary data remain sparse.

Importantly, LBM is not just comprised of skeletal muscle but also includes the heart, liver, bones and other tissues. Additionally, fat mass has a lean component, too[7]. As such, a decrease in LBM does not exclusively reflect changes in muscle mass but a diverse number of tissues including fat. Thus, distinguishing the effect of anti-obesity medications (AOMs) on muscle and other lean tissues in the absence of a direct measurement of muscle mass is challenging. Unfortunately, few clinical trials have directly assessed muscle after pharmacological weight loss, with only recently the first post-hoc reports emerging[8]. No clinical trial has examined changes in muscle function with AOMs. Even on the pre-clinical level, only two recent studies have reported changes to body composition together with muscle mass or function for a drug that is approved for obesity[9, 10]. Additionally, no study has explored how the new generation of incretin-based poly-agonists affects skeletal muscle.

Here we conducted a comprehensive assessment of changes to muscle mass and function with multiple incretin-based drugs and dosages in DIO mice. In the first study, we investigated the effect of a dual agonist (GLP-1R/GIPR) on changes in LBM, fat mass, and organ weight. In the second study, we examined the effect of GLP-1RA on muscle strength and endurance *in vivo*. In the third study, we explored whether in a known model of muscle wasting (i.e., immobilization), incretin-based therapies would accelerate muscle loss in obese mice. Finally, we conducted a proof-of-concept clinical study in patients with obesity (PWO), where we tested changes in BW, LBM, muscle mass and muscle strength before and after treatment with GLP-1RA.

To establish the basic contribution of muscle to changes in BW and body composition, we treated DIO mice via daily subcutaneous (s.c.) injections with tirzepatide (50 μg/kg or ∼10 nmol/kg) or a vehicle for 14 days. We decided to use a GLP-1R/GIPR dual agonist since they have proven more efficacious than GLP-1RA monotherapy in reducing BW in mice[11] and PWO [8, 12], and there is no data on muscle mass available. We found a robust reduction in BW of ∼35% from baseline (Fig. 1a-b), which was accompanied by a 73% reduction in body fat (Fig. 1c-d) and a 13% reduction in LBM (Fig. 1e-f). The reduction in LBM contributed ∼20% to the total weight loss. We collected five muscles of the lower limbs out of which only two showed a significant reduction in mass (Fig. 1g), each in the range of ∼10% (Fig. 1h). Importantly, since the loss of BW outpaced the decrease in muscle mass, the muscle weight per BW ratio (i.e., relative muscle mass) significantly improved in the extensor digitorum longus (EDL), soleus (SOL) and gastrocnemius (GSTN), and trended toward an improvement in the tibialis anterior (TA) and the quadriceps (QUAD) (Fig. 1i). In contrast, all white adipose tissue depots (WAT) were significantly reduced by ∼50-70%, with brown adipose tissue trending toward a significant reduction of ∼40% (Fig. 1j-k). Interestingly, liver mass numerically decreased by ∼20% (Fig. 1l). This decrease in liver mass is supported by results from a similar experiment, where we compared the effect of a GLP-1RA (semaglutide, 10 nmol/kg) and a GLP-1R/GIPR dual agonist (MAR709, 10nmol/kg) on BW, food intake and body composition. Here, we found a decrease in BW of 28 and 33% for semaglutide and MAR709, which was associated with lowered food intake, a reduction in body fat of ∼51 and 62%, a decline in LBM of ∼5 and 8%, and a significant decrease in liver mass of ∼20% (Supplemental Fig. 1a-e). Overall, this suggests that while LBM loss with incretin-based drugs can reach significance in DIO mice, the weight loss is primarily driven by a decrease in fat mass, resulting in an improvement in the muscle mass to BW ratio.

**Fig. 1:**
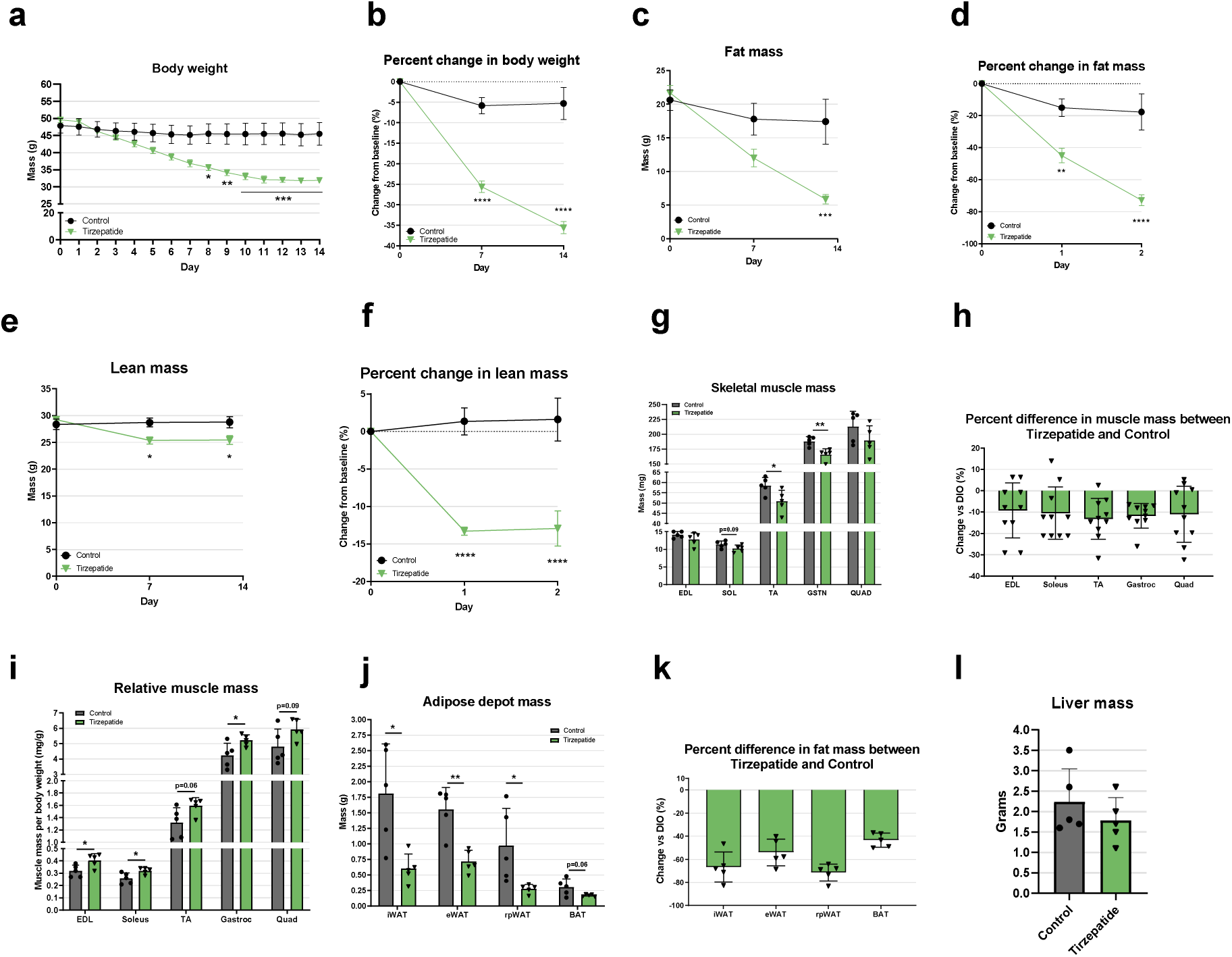
GLP-1R/GIPR dual agonism in DIO mice primarily reduces fat mass, and while LBM decreases significantly, the muscle mass to BW ratio improves. Body weight **(a-b)**, fat mass **(c-d)** and LBM **(e-f)** development over the course of the 14-day intervention. Group differences were determined via a two-way ANOVA and Sidak’s multiple comparison test. *, **, *** and **** correspond to p<0.05, p<0.01, p<0.001 and p<0.0001 when comparing tirzepatide to the control group. Absolute muscle mass **(g-h)** and relative muscle mass **(i)** as well as adipose tissue depots **(j-k)** and liver mass **(l)** at the end of the intervention. Group differences were determined via unpaired t-test between tirzepatide and the control group, with * and ** corresponding to p<0.05 and p<0.01, respectively. *n*=5 per group, unless otherwise denoted. For the muscle mass data in **(g)**, the average of both legs was calculated, while in **(h)** all individual values of the left and right leg are depicted together.

To test whether these results translate to improved mobility and performance in mice, our next experiment focused on skeletal muscle function with GLP-1RA. DIO mice received either semaglutide (40μg/kg or ∼10 nmol/kg) (Sema) or a vehicle injection for 28 days. To provide important context for the changes to muscle with pharmacological weight loss, we compared our obese groups to lean control mice on a regular chow diet. Absolute BW of the DIO vehicle group further increased over the course of the study, while the Sema group decreased and the lean group remained mostly weight stable (Fig. 2a). At 14 days, the difference between DIO and Sema was significant (Fig. 2a). Relative to the DIO vehicle group, Sema treatment resulted in a 22% decrease in BW on day 28 (Fig. 2b). In line with that, fat mass in the Sema group was significantly reduced compared to DIO starting at 14 days, with the difference on day 28 reaching 46% (Fig. 2c). This reflects the fact that absolute fat mass continued to increase in the DIO group over the course of 28 days, while it was reduced in the Sema group (Fig. 2d). In contrast, LBM was not statistically different between any of the groups at any time point (Fig. 2e). There was, however, a significant decrease in LBM in the Sema group from day 0 to day 28 (Fig. 2f). While the LBM decrease of 4% is much less substantial than the 46% decrease in fat mass, it still contributed roughly 32% of the total weight loss, owing to the fact that the mice carried almost twice as much LBM as fat at baseline (Fig. 2d & f). This supports the notion that incretin-based therapies do result in significant loss of LBM in obese mice.

**Fig. 2:**
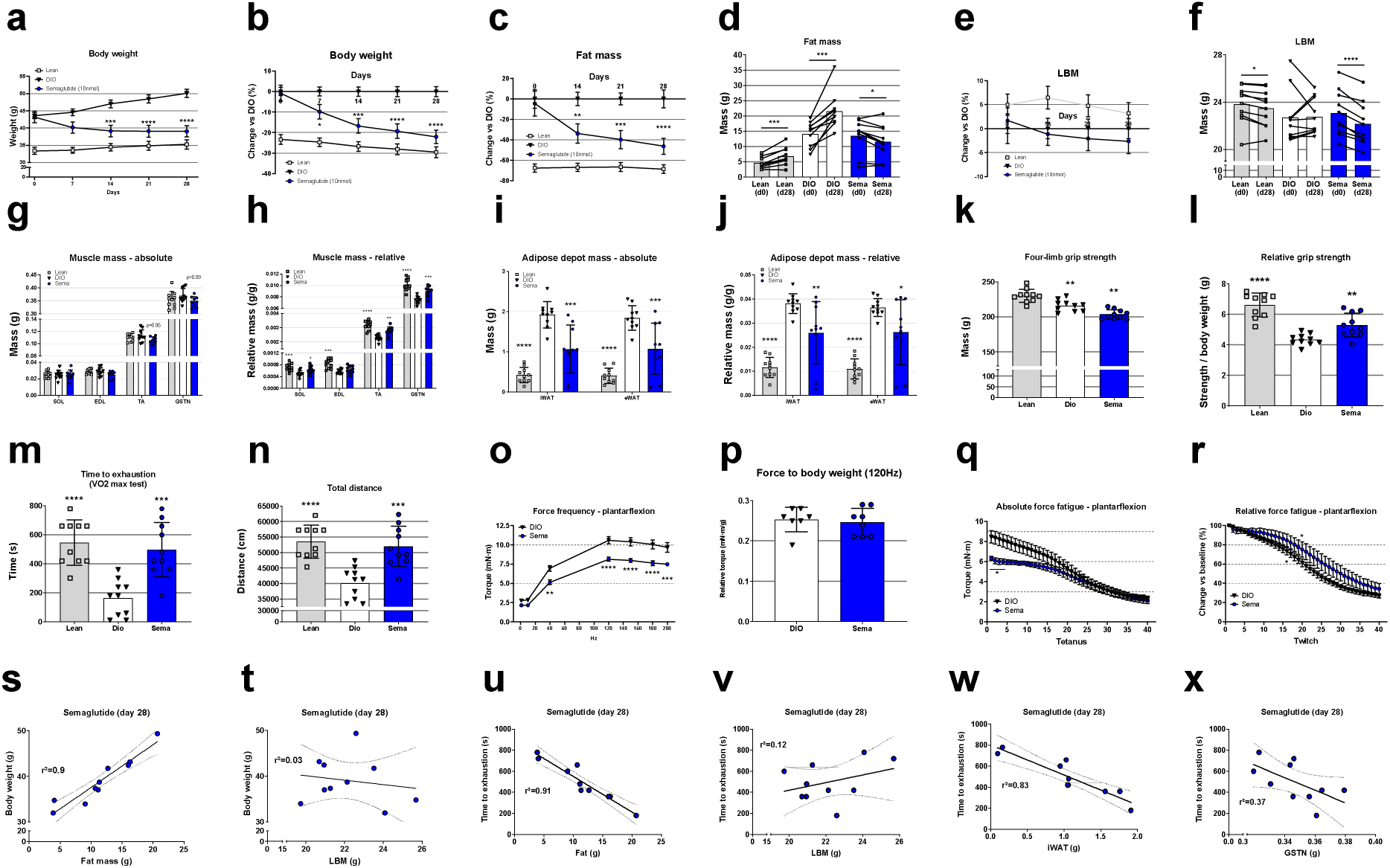
GLP-1RA reduces absolute muscle parameters but improves the muscle to BW ratio, leading to increased treadmill running performance. Absolute BW **(a)** and percent-change in BW vs DIO control **(b)** over the course of the 28-day intervention. Group differences were determined via two-way ANOVA and Tukey’s multiple comparison test. *** and **** denote p<0.001 and p<0.0001, respectively, for the comparison of Sema vs DIO. Percent-change in fat mass vs DIO control **(c)** and absolute values in fat mass at day 0 and day 28 of the intervention **(d)**. Group differences were determined via two-way ANOVA and Tukey’s multiple comparison test **(c)** or a paired t-test **(d)**. **, *** and **** denote p<0.01, p<0.001 and p<0.0001 for the comparison of Sema vs DIO **(c)**, while the asterisks in **(d)** denote the p-values for the before-and-after comparison within each group. **(e)** and **(f)** show the equivalent for LBM. **(g)** and **(h)** display absolute- and relative muscle mass, while **(i)** and **(j)** display absolute- and relative adipose tissue mass. Muscles were collected from both legs and adipose tissue from all respective sites, with the sum being displayed. Group differences were determined via one-way ANOVA and Dunnett’s multiple comparison test. *, **, *** and **** denote p<0.05, p<0.01, p<0.001 and p<0.0001 compared to DIO. Absolute grip strength **(k)** and grip strength relative to body weight **(l)** from all four limbs were collected. Time to exhaustion during a treadmill VO2 max test **(m)**, and total distance during the entirety of the running protocol **(n)** were recorded. For **(k-n)**, group differences were determined the same way as **(g-j)**. Force frequency **(o)**, force to body weight ratio (p), absolute force fatigue **(q)** and force fatigue relative to baseline **(r)** of the plantar flexors were assessed via *in vivo* contractility. Group differences were determined via two-way ANOVA and Sidak’s multiple comparison test **(o, q, r)** or an unpaired t-test **(p)**. *, **, *** and **** denote p<0.01, p<0.001 and p<0.0001 for the comparison of Sema vs DIO. Linear regression analysis was performed for BW vs fat mass **(s)**, BW vs LBM **(t)**, time to exhaustion vs fat mass **(u)**, time to exhaustion vs LBM **(v)**, and time to exhaustion vs iWAT **(w)** and GSTN **(x)**. All linear regression values were from day 28. Unless otherwise denoted, *n*=10 per group for the first intervention **(a-n**, and **s-x)**, and *n*=7-8 for the second intervention where in vivo contractility was tested **(o-r)**.

Since LBM is not exclusively comprised of skeletal muscle, we next investigated the major muscles of the lower limb. Interestingly, despite the decrease in LBM, we found only trends towards a reduction in absolute muscle mass with Sema (Fig. 2g). However, the amount of muscle relative to the BW of the animals improved significantly in the SOL, TA and GSTN with Sema compared to the DIO group (Fig. 2h). In accordance with the first experiment, this suggests that while overall muscle mass tended to decrease with GLP-1RA, muscle mass was spared relative to overall BW loss. Since fat mass was lost to a larger degree than LBM, we measured inguinal- and epididymal WAT to confirm this. Indeed, iWAT and eWAT decreased robustly by ∼45% and ∼41% in the Sema compared to the DIO group (Fig. 2i), resulting in a significantly lower adipose tissue to BW ratio for Sema (Fig. 2j).

To better understand how these changes in body composition and tissue weights affect muscle function and mobility, we examined grip strength, running performance, and *in vivo* muscle contractility. Similar to the trends in muscle mass, absolute grip strength decreased slightly with Sema compared to DIO (Fig. 2k), while relative grip strength improved (Fig. 2l). This improvement in the ratio of muscle mass and function to BW also resulted in a robust increase in time to exhaustion for the Sema mice during a VO2 max treadmill test (Fig. 2m). Indeed, the time to exhaustion and the total distance covered by the Sema mice was not just higher than the DIO mice but almost even with the lean control group (Fig. 2n). Maximal torque during a force frequency test of the plantar flexors followed a similar trend: absolute force at higher frequencies was decreased in Sema versus (vs) DIO mice (Fig. 2o) but relative torque (i.e., the torque to BW ratio) was unchanged (Fig. 2p). Since we found absolute strength parameters (grip strength, force frequency) to be slightly reduced but endurance readouts improved (treadmill performance), we next tested how fatigue is affected during a repeated tetanus-torque test. In alignment with the previous results, the initial peak torque was significantly higher in the DIO compared to the Sema group (Fig. 2q). However, the decline in torque began significantly later in the Sema group, indicating an improvement in fatigue resistance (Fig. 2r). This reinforces our finding that while absolute muscle mass and strength appear to mildly decrease with GLP-1RA, this effect is offset by a greater loss of adipose tissue, resulting in a higher relative muscle mass and increased mobility. To confirm that changes in fat mass and AT are more predictive for BW loss and running performance than LBM and muscle, we also conducted a sub-cohort linear regression analysis for these variables in the Sema group. We found that the final BW of the Sema mice on day 28 correlated remarkably with fat mass (r^2^=0.9) (Fig. 2s) but not with LBM (r^2^=0.03) (Fig. 2t). Similarly, time to exhaustion was negatively associated with fat mass (r^2^=0.91) (Fig. 2u) and iWAT (r^2^=0.83) (Fig. 2w), but not LBM (r^2^=0.12) (Fig. 2v). In fact, better running performance appeared mildly associated with smaller GSTN mass (r^2^=0.37) (Fig. 2x).

Next, we wanted to test how the use of incretin mimetics affects skeletal muscle during a known stimulus for wasting. We chose immobilization as it has been shown to result in rapid and substantial disuse atrophy in rodents and humans[13, 14]. Additionally, repeated periods of immobilization have been proposed as a major risk factor for sarcopenia[15]. Since most of our muscle-related results in DIO mice under ambulant conditions could be explained through weight loss, we hypothesized that the more extreme combination of AOMs and immobilization may reveal more subtle, negative effects of incretin-based therapies. We kept obese mice either untreated (DIO), unilaterally casted using a 3D printed cast described by Moore and colleagues[16] and vehicle injected (Cast), GLP-1RA treated (80μg/kg or ∼20nmol/kg semaglutide) and unilaterally casted (Sema) or GLP-1R/glucagon receptor (GCGR) dual-agonist treated (80μg/kg or ∼20nmol/kg survodutide) and unilaterally casted (Survo). We chose a GLP-1R/GCGR dual-agonist in addition to GLP-1RA monotherapy, because pre-clinical data reported a significant reduction of LBM at higher doses[17] and a phase 2a study found a reduction in circulating amino acids[18]. Based on glucagon’s role in liver metabolism and its ability to suppress muscle protein synthesis[19] this raises the concern that GCGRA could directly impact muscle loss by promoting the liberation of amino acids from skeletal muscle and conversion to glucose in the liver[20, 21]. Additionally, data suggest that dual- and triple agonists containing a GCGRA are superior to GLP-1RA monotherapy for weight loss [17, 22], highlighting their future potential for clinical practice. To directly compare pharmacological with physiological weight loss, we also included a calorie-restricted group (PairFed) whose food intake was matched to the Sema group (Fig. 3a).

**Fig. 3:**
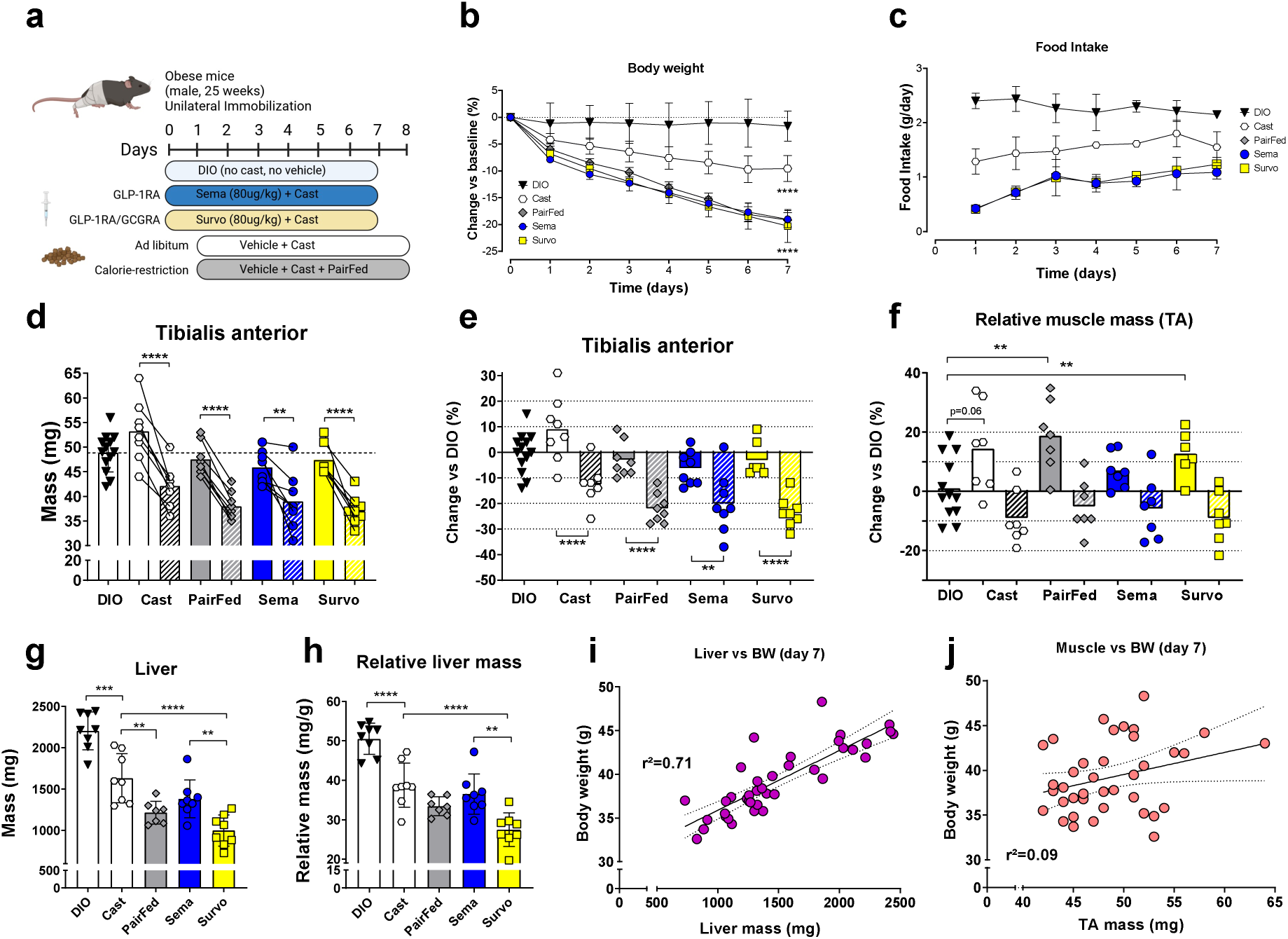
Pharmacological and physiological weight loss result in similar reductions of body weight and muscle mass during immobilization. Graphical representation of the immobilization experiment **(a)**. Change in BW **(b)** and food intake **(c)** over the course of the 7-day intervention. Group differences for **(b)** were determined via two-way ANOVA and Tukey’s multiple comparison test. Significant differences compared to the DIO control on day 7 are depicted as **** corresponding to p<0.0001. Differences in food intake were not statistically assessed. Absolute TA mass **(d)**, change in TA mass vs baseline **(e)** and TA mass relative to the BW of the animals **(f)** were collected on day 7. Within-group differences between the control leg and the casted leg **(d-e)** were determined via a paired t-test, with ** and **** denoting p<0.01 and p<0.0001, respectively. Across-group differences between the DIO group and all other interventions were determined via a one-way ANOVA and Dunnett’s multiple comparison test **(f)**, with ** denoting p<0.01 when comparing the indicated leg to the DIO control. Absolute and relative liver mass **(g-h)** were collected on day 7. Group differences were determined via a one-way ANOVA and Tukey’s multiple comparison test. **, *** and ****, denote p<0.01, p<0.001 and p<0.0001 between the indicated groups, respectively. Linear regression analysis was performed for BW vs liver mass **(i)** and BW vs TA mass **(j)**. All linear regression values were from day 7 of the intervention. *n*=8 per group, unless otherwise denoted. For the muscle mass data of the DIO control group, left and right leg data were clustered as one group.

After 7 days, the group with a cast and *ad libitum* feeding lost ∼10% BW, while the PairFed, Sema, and Survo groups all lost ∼20% BW (Fig. 3b). That the cast control group lost significant weight despite *ad libitum* food access is in line with our previous observations in hindlimb-suspended rats[13]. The food intake over the course of the intervention mirrored the change in BW of all groups (Fig. 3c). Casting decreased TA muscle mass in all treatment groups compared to the control leg (Fig. 3d). Interestingly, compared to the untreated DIO group, the control leg of the casted mice with *ad libitum* food intake showed a numerical increase of ∼9% (Fig. 3e). This suggests that the ambulant leg was relied on and loaded more, resulting in mild compensatory hypertrophy. We did not observe a net increase in control TA mass in any other treatment groups, likely owing to the more robust weight loss. Indeed, when looking at the amount of muscle mass relative to the BW, all casted groups showed a numerical increase in the control leg compared to untreated DIO mice (Fig. 3f). The increase in the control leg of the Cast group showed a trend, and the improvements in the PairFed and the Survo group were even significant (Fig. 3f). While the absolute decrease in TA mass of the casted leg ranged from -13% (Cast) to -24% (Survo) compared to untreated DIO mice (Fig. 3e), the relative decrease (i.e., accounting for BW change) was just between -5% (PairFed) to –9% (Survo) (Fig. 3f).

Since our data from Fig. 1 suggested a robust involvement of the liver, we also investigated changes in liver mass after combined immobilization and incretin mimetic treatment. We found that liver mass decreased considerably in all treatment groups, -26% in Cast, 45% in PairFed, 37% in Sema, and 55% in Survo (Fig. 3g). The fact that Survo achieved a significantly greater decrease in liver mass compared to Sema, despite similar weight loss, can likely be explained by the GCGRA effects on the liver and is in agreement with phase 2 clinical data that showed a robust reduction in liver fat of patients living with metabolic dysfunction-associated steatohepatitis (MASH) [23]. As such, a previously significant difference in liver mass between Cast and PairFed disappeared when we normalized to BW, but the difference between Sema and Survo persisted (Fig. 3h). Further support for the strong involvement of the liver in regulating BW with calorie restriction and incretin-based therapies was found when we performed a linear regression analysis that included all groups in our study. While liver mass correlated tightly with BW (r^2^=0.71) (Fig. 3i), muscle mass in the uncasted TA was found to be a remarkably poor predictor of BW (r^2^=0.09) (Fig. 3j). Taken together, this suggests that when matched for calorie intake, pharmacological and physiological interventions result in comparable weight loss, with both having relatively modest effects on muscle mass in DIO mice. Consistent with this, immobilization caused a reduction in TA mass that was impacted by the extent of weight loss but not by the type of weight loss (i.e., physiological vs pharmacological).

To explore how our findings in DIO mice translate to the human condition, we performed a proof-of-concept clinical trial in patients with obesity and diabetes. Some endpoints of this trial such as body weight and body composition have been described previously [24]. However, here we report for the first time how GLP-1RA treatment affected muscle mass and function in our patients. Briefly, PWO and diabetes were escalated from 0.25mg semaglutide s.c. weekly to 1mg within the first four weeks of the trial (where tolerated) before maintaining this dose for another 8 weeks. BW, fat mass, and LBM all decreased significantly over the course of the 12 weeks [24]. However, the contribution of fat mass to weight loss was more than twice as high as LBM (70 to 30%, respectively). This contribution of LBM is less than what the STEP 1 trial found for semaglutide[2], but more than what the SURMOUNT-1 trial recently reported for tirzepatide[25]. Accordingly, we found that the fat to BW ratio significantly decreased in our patients (Fig. 4a), while the LBM to BW ratio significantly increased (Fig. 4b), resulting in favorable changes to body composition. In line with our animal data, absolute measures of muscle size decreased (Fig. 4c) while relative muscle size was not negatively affected (Fig. 4d). Interestingly, despite the significant reduction in cross sectional area (CSA) of the vastus lateralis (VL), neither absolute muscle strength (MVC of the knee extensors) (Fig. 4e) nor relative muscle strength decreased (Fig. 4f). To our knowledge, this is the first time that direct measurements of muscle mass and strength were reported alongside each other for a clinical trial of incretin-based weight loss. The apparent discrepancy between muscle size and function could indicate that the reduction in CSA may not be entirely comprised of contractile proteins but also a decrease in other cellular components, like substrates. This would be in line with recent reports of decreased intramuscular fat after prolonged GLP-1RA/GIPRA treatment [8]. Similar to the lack of change in MVC, neither absolute (Fig. 4k), nor relative, hand grip strength (Fig. 4l) were negatively affected by our intervention.

**Fig. 4:**
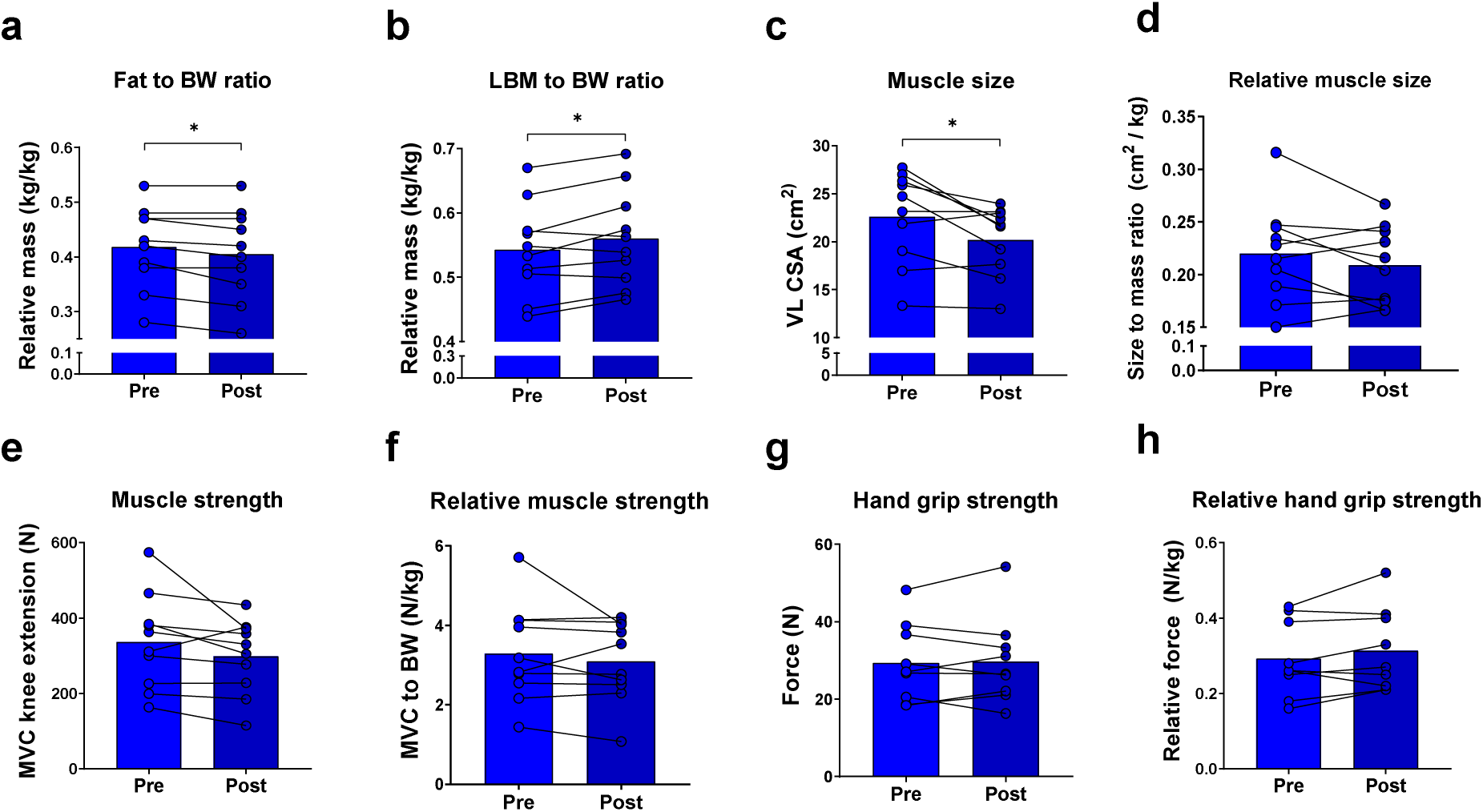
GLP-1RA treatment in patients results in a preferred reduction of body fat over LBM, and a relative preservation of muscle strength despite a modest decrease in muscle size. Decrease in fat to BW ratio **(a)** and increase in LBM to BW ratio **(b)** after 12 weeks of GLP-1RA treatment. Absolute muscle size **(c)** and relative muscle size **(d)** as well as absolute muscle strength **(e)** and relative muscle strength **(f)**, and absolute as well as relative hand grip strength **(g-h)**. Group differences were assessed via paired t-test, with * and **** denoting p<0.05 and p<0.0001, respectively. *n*=10 per group, unless otherwise denoted.

## Discussion

Body composition data from large-scale clinical trials have sparked a concern for muscle wasting with incretin-based therapies. Due to the widespread use of these drugs, such complications would have severe public health ramifications and thus have been the focus of various reviews[4, 7]. Despite these growing concerns, primary data on muscle wasting with incretin treatments remain sparse No clinical study has yet assessed muscle mass and function in obese patients treated with weight loss drugs for a prolonged period of time. At the pre-clinical level, two studies have shown that myostatin/activin A inhibition can prevent muscle loss in pre-clinical models of incretin-treated obesity [9, 26]. However, neither of the studies included a lean control- or pair-fed group to explore whether the muscle mass changes with incretins were disproportionate or a simple function of weight loss. Similarly, a comprehensive assessment of muscle function and strength after AOM treatment has been missing, particularly in the clinical context.

We found support for the notion that muscle mass decreases as part of drug-induced weight loss. However, we did not find any evidence that this muscle loss was disproportionate or pathological. Across several pre-clinical scenarios and compounds, we found a consistent absolute decrease in muscle mass but a relative improvement in muscle to BW ratio. Even when challenged with a high dose of a GLP-1RA or a GLP-1R/GCGR dual agonist during immobilization, changes in muscle mass were remarkably similar to physiological weight loss through calorie restriction. While maintaining muscle mass as the largest depot of glucose in the body has multiple metabolic advantages to offer, preserving muscle function might be even more important clinically [27]. This is particularly true in the context of aging, since strength is lost faster than muscle mass [28, 29]. However, we found that the decrease in muscle strength was similar to the trend in muscle mass, with both conserved relative to BW. This resulted in a favorable power to BW ratio in GLP-1RA-treated mice, which led to substantially improved treadmill performance that was comparable to lean animals.

In our GLP-1RA clinical trial, we observed similar tendencies. While LBM decreased significantly, a much larger portion of the weight loss was due to decreased fat mass. Even though muscle size decreased significantly, maximum knee extension, or hand grip strength remained unchanged. While our trial was only designed as a proof-of-concept and cannot provide conclusive evidence, it was the first to directly assess muscle function in obese patients treated with incretin-mimetics. However, recent large-scale clinical trials such as the STEP 9 in obese patients with osteoarthritis support the notion that the disproportionately larger loss of fat mass than LBM results in improved mobility and physical function [30]. Similar findings were reported by the SLIM LIVER study, where trends towards improved gait speed were found despite a decrease in psoas muscle volume[31]. Remarkably, this was found despite the patients suffering from a combination of human immunodeficiency virus (HIV) and MASH. Together with our data, these findings highlight that for PWO and related metabolic diseases, reducing BW and fat mass is paramount to improving health and mobility, even if it comes at the expense of a mild loss of LBM and muscle.

Importantly, our results also underscore that whole-body LBM and muscle mass should not be conflated. While this important concept has been pointed out by colleagues in the context of AOMs [7, 32], the contribution of organs outside of skeletal muscle to LBM changes with incretin-based therapies is still poorly understood. By comparing changes in liver and muscle mass simultaneously, we found that the relative change in liver mass is much more pronounced during weight loss in obese mice. Importantly, we found this to be the case with various AOMs (GLP-1RA, GLP-1RA/GIPRA, GLP-1RA/GCGRA) as well as physiological weight loss (i.e., calorie restriction), with the effect being most pronounced for GLP-1RA/GCGRA. The fact that liver mass is so robustly affected by weight change raises another important issue for the interpretation of body composition data, highlighting that changes to LBM cannot be extrapolated to muscle mass or function. This is particularly important given that the tool routinely used to assess body composition in clinics, dual x-ray absorptiometry (DXA), cannot distinguish between LBM and fat that resides within predominantly lean tissues. In other words, a decrease in intra-hepatic or intra-muscular substrate content (i.e., triglycerides or glycogen) cannot be distinguished by DXA, and will inevitably be quantified as a decrease in LBM. Since we found a strong decrease in liver mass in our pre-clinical cohorts, and clinical trials have shown a robust decrease in liver fat with incretin-based therapies [23, 33, 34] that cannot be captured via DXA, this could be another reason why some clinical trials have found more LBM loss than anticipated. Notwithstanding these methodological limitations and confounders, it is also important to point out that unlike the STEP 1 trial, the majority of clinical studies with incretin-based treatments did not find a disproportionate contribution of LBM to weight loss, but were remarkably close to the “quarter FFM rule”[7].

As such, current evidence provided by our study and the literature does not support the concern for an accelerated loss of muscle mass, function, or mobility with incretin-based treatments in obese mice or patients. Nevertheless, future work will have to assess muscle mass and function in incretin-treated patients who have additional comorbidities, particularly conditions known to cause muscle wasting (i.e., sarcopenia, cachexia or heart disease). Similarly, even in the absence of a pathological effect of incretins on muscle, improved maintenance of muscle mass during weight loss may still confer metabolic and functional advantages that result in higher resilience towards age-related diseases. Therefore, finding therapeutic solutions targeting muscle mass and function continues to be of high clinical relevance. Lastly, long-term adherence to incretin-based drugs is known to be poor, with over half of the patients discontinuing treatment within one year[35], raising the concern of repeated weight loss- and regain, which may result in deteriorating body composition[36, 37]. Future studies will need to address whether AOMs exacerbate these effects and explore the role of skeletal muscle in preventing it.

## Data Availability

All data produced in the present study are available upon reasonable request to the authors.

## Data availability

All data used for the statistical analysis are available in the data source file, along with the GraphPad Prism-derived report on the statistical analysis.

## Acknowledgements

We would like to thank Dr Laura Moore for sharing the 3D cast computer aided design files. Body composition measurements in Fig. 1 were performed through the University of Pennsylvania Rodent Metabolic Phenotyping Core (RRID:SCR_022427), supported in part by NIH grant S10-OD025098.

## Methods

### Ethics

All animal studies were approved by their respective authorities as listed for each individual experiment in the sections below. The proof-of-concept clinical trial prospectively received ethical approval from local authorities (East of England, Essex Research Ethics Committee) and was later registered on clinicaltrials.gov (NCT05606471) [24].

### Sub chronic DIO study with GLP-1RA/GIPRA dual agonist treatment

All mouse experiments in Fig. 1 were reviewed and approved by the University of Pennsylvania IACUC in accordance with National Institutes of Health guidelines. Male, diet-induced obese mice (18 weeks of age) on a C57Bl/6J background were purchased from Jackson Laboratory (#380050) and maintained on a high fat diet (60 kcal% from fat; Research Diets D12492) for the duration of the experiment. Mice were group housed and had *ad libitum* access to food. Tirzepatide (Peptide Sciences) or an equal volume of vehicle control (0.9% saline) were administered s.c. at 50 μg/kg once daily for 14 days. Body weight was measured daily and body composition was assessed at days 0, 7, and 14 using an EchoMRI body composition analyzer (EchoMRI, Houston, TX). Mice were euthanized on day 14 and fat pads (eWAT, iWAT, rpWAT, BAT), hindlimb muscles (TA, SOL, EDL, GSTN, QUAD), and liver were collected and weighed.

For the sub chronic study mentioned in Supplemental Fig. 1, all experiments were performed in accordance with the animal protection law of the European Union after permission by the government of Upper Bavaria, Germany. 54-week-old C57BL6/J male mice were double-housed and fed ad libitum with a high-fat diet (58% fat, D12331, Research Diets, New Brunswick, USA). Fat and LBM were measured via nuclear magnetic resonance technology (EchoMRI, Houston, USA). Semaglutide and MAR709 were provided by Novo Nordisk (Indianapolis, USA). Animals were injected daily (s.c.) with the drugs or vehicle. On day 14, the mice were sacrificed and liver weight recorded.

### Sub chronic DIO study that assessed muscle function

Adult male C57BL/6J mice at 18 weeks old were purchased from JAX (Bar Harbor, ME, USA) after being fed for 12 weeks with either 60% high-fat diet (HFD; Research Diets# D12492i; DIO mice, *n*=20) or normal chow diet (NC; SAFE A04i ; NC mice, *n*=10). After receiving the animals, the preconditioned DIO mice and lean NC mice were acclimated to Biomeostasis’ animal facility for two weeks, followed by one week of daily habitation to s.c. injections (vehicle). Throughout the study, mice were group housed (*n*=2/cage) and allowed *ad libitum* access to water and HFD or NC, and maintained in Biomeostasis’ animal facility at ambient temperature (22°C) and in a humidity (40-60 %) controlled room on a 12-hour light-dark cycle. Animal handling and procedures were conducted under the protocols and/or guidelines approved by the Institutional Animal Care and Use Committee (IACUC) and the local ethics committee (Marseille, APAFIS #49719). Mice received daily s.c. injections of vehicle (saline) or 40μg/kg semaglutide (Peptide Sciences, Lot# VIM982031106-3) for 4 weeks. On D0 (prior to the first dosing), D14, D21 and D28, body composition was measured using a TD-NMR minispec Analyzer (LF90II, Bruker, Germany).

### Grip strength

On D28, the combined forelimb and hindlimb grip strength was measured on a grid assembly connected to a grip strength meter (BIOSEB, BIO-GS3). Briefly, the mouse is placed over the top of the grid so that both its front paws and hind paws can grip the grid. Torso was kept parallel to the grid and the mouse was pulled back steadily until the grip was released entirely from the grid. Upon release, maximal grip strength was automatically recorded by the acquisition software (BIO-CIS; BIOSEB, France). The procedure was repeated 3 times per animal with 5 minutes intervals between measurements. Grip strength was calculated as the mean of the 3 repeated measurements.

### Treadmill running

On D21, each mouse was allowed to acclimate to a metabolic treadmill for 10 minutes at 10 m/min with no incline (0°) in order to familiarize the animals with the treadmill before the exercise test. On D28, each mouse ran on a one-lane metabolic treadmill (OxyletPro - Panlab LE 8708) connected to a gas analyzer (OxyletPro – Panlab LE405) for VO2 assessment. Each mouse was allowed to acclimate to the treadmill for 1h to obtain baseline values. The treadmill was then switched on for 30 min of light running at 10 m/min with no incline (0°). Subsequently, VO2 max was assessed during a progressive test of 2 min segments where either speed or inclination was increased: 15 m/min at 0°, 20 m/min at 0°, 20 m/min at 5°, 20 m/min at 10°, 20 m/min at 15°, 20 m/min at 20°, 20 m/min at 25°, 25 m/min at 25° and 30 m/min at 25°. The VO2 max test was stopped when a mouse remained on the shock pad for 10s. VO2 max was calculated as the mean VO2 during the last 3 min of the protocol and “time to exhaustion” as the completed time on the treadmill per mouse.

### *In vivo* contractility

To avoid interference between the assessment of muscle mass, running performance and contractility, muscle force was tested on a separate set of animals that underwent the same drug intervention as described above but without the grip strength and treadmill running. No NC animals were included in this experiment. On D28, *in vivo* muscle strength of the right leg was tested using an Aurora Scientific System (1305A in vivo/in situ/in vitro muscle test system - 5N), similar to what has been described previously[38, 39]. Briefly, mice were anaesthetized (isofluorane/oxygen, 1 l/min) and the upper portion of the hind limb was held in place, with the right paw fixed to the pedal of the servomotor system (301C, Aurora Scientific Inc., Aurora, Canada) such that only dorsiflexion and plantar flexion of the ankle were possible. Plantarflexion was stimulated through insertion of needle electrodes adjacent to the sciatic nerve). The following protocols were used: for the force frequency assay, a series of stimulations were performed at increasing frequencies (0.2 ms pulse, 500 ms train duration): 1, 10, 40, 120, 140, 160, 180 and 200 Hz. For the fatigue assay, a series of 40 stimulations at 120 Hz (tetanus stimulation frequency) were performed. On D28, the mice were fasted for 4h before being collected, and the weight of the GSTN, TA, EDL, SOL, iWAT and eWAT recorded.

### Immobilization experiment

All immobilization experiments were approved by the University of California Davis Institutional Animal Care and Use Committee under Protocol #23367. Forty male Black 6 DIO mice (age 19 weeks) were obtained from Jackson Laboratories and housed 4 per cage with 12h light/dark cycles and ad libitum access to food and water. The right limbs of mice were immobilized as described by Moore and colleagues[16]. Briefly, 3D printed casts were placed over the right knee and ankle joint and held in place using 10lb wire. Both joints were maintained in a neutral position over the next seven days. Over the course of the study, none of the animals removed their casts. Starting at the time of immobilization, animals received subcutaneous injections of 80μg/kg or ∼20nmol/kg semaglutide (Sema), 80μg/kg or ∼20nmol/kg survodutide (Survo), or vehicle. A separate group of true control DIO mice were maintained without casting or injection.

Food was weighed daily, and total consumption was divided by the number of mice per cage to determine average food intake. The average food intake of the Sema group was then weighed out and provided to a separate group of one-day delayed casted, vehicle injected and single-housed mice (Pairfed). BW of all animals were tracked daily for the 7 days of study. At collection, the casts were removed and the tibialis anterior muscle removed, weighed, and frozen in liquid nitrogen. Livers were removed and weighed to estimate lean mass loss from other metabolic tissues.

### Pilot clinical trial

Participant recruitment, eligibility criteria and study design have been described in prior publication reporting changes to body weight, -composition, and metabolic effects [24]. Data presented here are novel unpublished data on muscle mass and function following semaglutide treatment in PWO and type 2 diabetes (T2D). Briefly, adults aged between 18-75 years that had been diagnosed with T2D and a BMI between 27-50kg*m^-2^ were recruited into the study. Participants followed the study schedule previously described and therefore received once weekly subcutaneous semaglutide (Novo Nordisk) for 12 weeks, commenced at 0.25 mg and escalated every two weeks to 1 mg. This rapid dose escalation was performed to maximise exposure to the full dose of semaglutide and facilitate the fairest comparison between the interventions. Adverse event data were collected and if participants experienced significant side effects, dose escalation was delayed as would in usual clinical practice.

Each participant in this group was supplied with dietary advice focussed on portion size reduction, avoidance of food with high-fat content, keeping well hydrated, and increasing awareness of satiety. This advice was provided in a written format along with the dosing schedule of semaglutide. Participants were guided through administration of the first dose and then self-administered the remaining doses. They were instructed to document each dose they administered, to ensure they followed the study’s dosing schedule.

### Clinical assessment of muscle size and strength

VL muscle structure was examined using B-mode ultrasonography while participants were laid in the supine position. The belly of the VL was identified by measuring its entire length and noting the midpoint of this line. Muscle cross-sectional area (CSA) was determined along the mid-line using panoramic image rendering between the medial and lateral borders. Muscle thickness was also measured along the longitudinal axis of the muscle by aligning the ultrasound probe along the fascicle plane at the VL midpoint. At least three images of each parameter were saved for subsequent analysis which was performed using Image J (National Institute of Health) software. MVC was assessed in an isometric dynamometer, with the leg fixed in a 90 degree flexion. The participant was asked to contract this leg as hard as possible against a force transducer, which simultaneously measures the force generated and displays this on a connected monitor. This is in contrast to 1-repetition maximum (1RM) testing which involves repeated attempts at moving a selected weight which increases following each successful attempt. Handgrip strength was assessed using a digital handgrip dynamometer (Grip D 5401, Takei Scientific Instruments, Japan).

### Statistics

Statistical analyses were performed using GraphPad Prism software v.8 (La Jolla, USA). Data in the main figures is reported as individual data points ± SD except for body weight and body composition data of animal experiments, which is reported a mean ± SEM for visibility. Statistical tests were according to the research question and included unpaired- and paired t-tests, one-way and two-way ANOVA, together with post-hoc analyses via Dunnett’s, Tukey’s and Sidak’s multiple comparison tests, respectively. For linear regression analyses, the line of best fit was calculated and displayed together with the 95% confidence intervals and the r² value for goodness of fit. Significance was determined as p<0.05 or smaller, with asterisk * corresponding to p<0.05, ** to p<0.01, *** to p<0.001 and **** to p<0.0001. The underlying statistical test for each comparison that resulted in a significant difference is listed in the descriptions corresponding to the figure.

## Supplemental Figures

**Supplemental Fig. 1:**
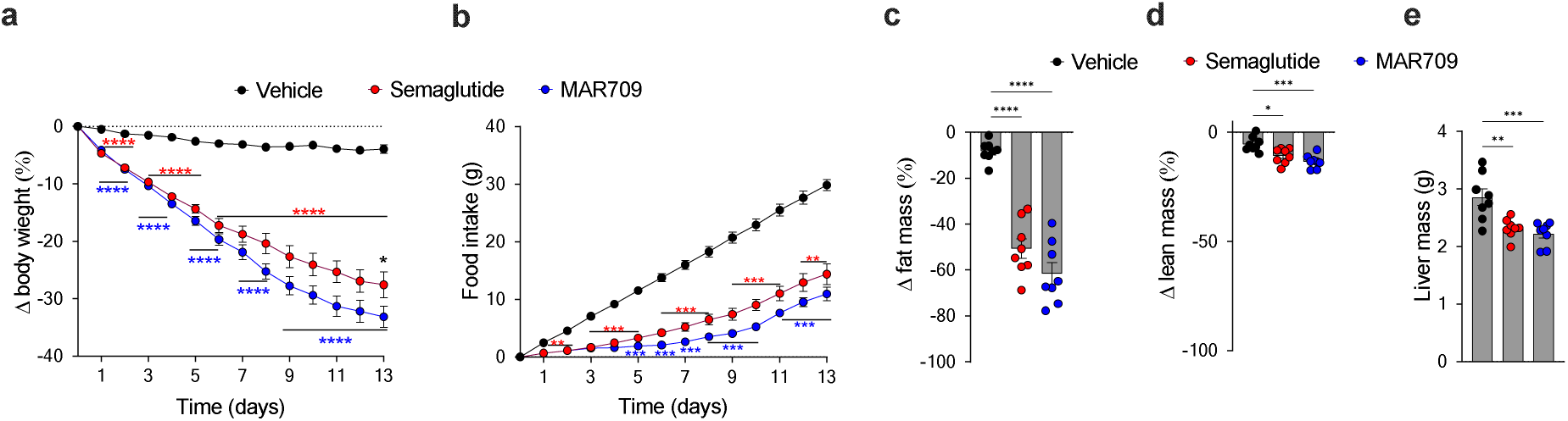
GLP-1R/GIPR dual agonism robustly decreases liver mass as part of a reduction in body weight, fat mass, and LBM. Body weight **(a)** and cumulative food intake **(b)** of 54-wk old male DIO C57BL/6J wildtype mice treated daily with either vehicle or 10 nmol/kg of either semaglutide or MAR709 (n=8 each group). Body fat **(c)** lean tissue mass **(d)** and liver mass at study day 14 (n=8 each group). Data in panel **a** and **b** were analyzed by repeated measures 2-way ANOVA with Bonferroni’s post-hoc test for comparison of individual time points. Data in panel **c, d and e** were analyzed using 1-way ANOVA with Tukey post-hoc test. t. Cumulative food intake in panel **b** (n=8 each group) was assessed per cage in double-housed mice. Data represent mean ± SEM; asterisks indicate * p<0.05; ** p<0.01 and *** p<0.001. Red asterisks in panel **a** and **b** correspond to the comparison of semaglutide vs vehicle, while blue asterisks correspond to MAR709 vs vehicle while black asterisks correspond to semaglutide vs MAR709.

## References

1. Federation, W.O., World Obesity Atlas 2025. London: World Obesity Federation, 2025, 2025.

2. Wilding, J.P.H., et al., Once-Weekly Semaglutide in Adults with Overweight or Obesity. N Engl J Med, 2021. 384(11): p. 989–1002.

3. Jastreboff, A.M., et al., Tirzepatide Once Weekly for the Treatment of Obesity. N Engl J Med, 2022. 387(3): p. 205–216.

4. Prado, C.M., et al., Muscle matters: the effects of medically induced weight loss on skeletal muscle. Lancet Diabetes Endocrinol, 2024. 12(11): p. 785–787.

5. Prentice, A.M., et al., Physiological responses to slimming. Proc Nutr Soc, 1991. 50(2): p. 441–58.

6. Arnold, C., After obesity drugs’ success, companies rush to preserve skeletal muscle. Nat Biotechnol, 2024. 42(3): p. 351–353.

7. Tinsley, G.M. and S.B. Heymsfield, Fundamental Body Composition Principles Provide Context for Fat-Free and Skeletal Muscle Loss With GLP-1 RA Treatments. J Endocr Soc, 2024. 8(11): p. bvae164.

8. Sattar, N., et al., Tirzepatide and muscle composition changes in people with type 2 diabetes (SURPASS-3 MRI): a post-hoc analysis of a randomised, open-label, parallel-group, phase 3 trial. 2025.

9. Nunn, E., et al., Antibody blockade of activin type II receptors preserves skeletal muscle mass and enhances fat loss during GLP-1 receptor agonism. Mol Metab, 2024. 80: p. 101880.

10. Xiang, J., et al., GLP-1RA Liraglutide and Semaglutide Improves Obesity-Induced Muscle Atrophy via SIRT1 Pathway. Diabetes Metab Syndr Obes, 2023. 16: p. 2433–2446.

11. Knerr, P.J., et al., Next generation GLP-1/GIP/glucagon triple agonists normalize body weight in obese mice. Mol Metab, 2022. 63: p. 101533.

12. Rodriguez, P.J., et al., Semaglutide vs Tirzepatide for Weight Loss in Adults With Overweight or Obesity. JAMA Intern Med, 2024. 184(9): p. 1056–1064.

13. Baehr, L.M., et al., Muscle-specific and age-related changes in protein synthesis and protein degradation in response to hindlimb unloading in rats. Journal of Applied Physiology, 2017. 122: p. 1336–1350.

14. Wall, B.T., et al., Substantial skeletal muscle loss occurs during only 5 days of disuse. Acta Physiol (Oxf), 2014. 210(3): p. 600–11.

15. Wall, B.T., M.L. Dirks, and L.J.C. van Loon, Skeletal muscle atrophy during short-term disuse: implications for age-related sarcopenia. Ageing research reviews, 2013.

16. Moore, L.K., et al., A novel mouse model of hindlimb joint contracture with 3D-printed casts. J Orthop Res, 2022. 40(12): p. 2865–2872.

17. Zimmermann, T., et al., BI 45CS0C: Discovery and preclinical pharmacology of a novel GCGR/GLP-1R dual agonist with robust anti-obesity efficacy. Mol Metab, 2022. 66: p. 101633.

18. Klein, T., R. Augustin, and A.M. Hennige, Perspectives in weight control in diabetes - Survodutide. Diabetes Res Clin Pract, 2024. 207: p. 110779.

19. Preedy, V.R. and P.J. Garlick, The effect of glucagon administration on protein synthesis in skeletal muscles, heart and liver in vivo. Biochem J, 1985. 228(3): p. 575–81.

20. James, H., et al., The Effect of Glucagon on Protein Catabolism During Insulin Deficiency: Exchange of Amino Acids Across Skeletal Muscle and the Splanchnic Bed. Diabetes, 2022. 71(8): p. 1636–1648.

21. Okun, J.G., et al., Liver alanine catabolism promotes skeletal muscle atrophy and hyperglycaemia in type 2 diabetes. Nat Metab, 2021. 3(3): p. 394–409.

22. Jastreboff, A.M., et al., Triple-Hormone-Receptor Agonist Retatrutide for Obesity - A Phase 2 Trial. N Engl J Med, 2023. 38G(6): p. 514–526.

23. Sanyal, A.J., et al., A Phase 2 Randomized Trial of Survodutide in MASH and Fibrosis. N Engl J Med, 2024. 3G1(4): p. 311–319.

24. Anyiam, O., et al., Metabolic effects of very-low calorie diet, Semaglutide, or combination of the two, in individuals with type 2 diabetes mellitus. Clin Nutr, 2024. 43(8): p. 1907–1913.

25. Look, M., et al., Body composition changes during weight reduction with tirzepatide in the SURMOUNT-1 study of adults with obesity or overweight. Diabetes Obes Metab, 2025. 27(5): p. 2720–2729.

26. Mastaitis, J.W., et al., GDF8 and activin A blockade protects against GLP-1-induced muscle loss while enhancing fat loss in obese male mice and non-human primates. Nat Commun, 2025. 16(1): p. 4377.

27. Li, R., et al., Associations of Muscle Mass and Strength with All-Cause Mortality among US Older Adults. Med Sci Sports Exerc, 2018. 50(3): p. 458–467.

28. Langer, H.T., et al., Commentaries on viewpoint: rejuvenation of the term sarcopenia. 2019.

29. Goodpaster, B.H., et al., The loss of skeletal muscle strength, mass, and quality in older adults: the health, aging and body composition study. J Gerontol A Biol Sci Med Sci, 2006. 61(10): p. 1059–64.

30. Bliddal, H., et al., Once-Weekly Semaglutide in Persons with Obesity and Knee Osteoarthritis. N Engl J Med, 2024. 3G1(17): p. 1573–1583.

31. Ditzenberger, G.L., et al., Effects of Semaglutide on Muscle Structure and Function in the SLIM LIVER Study. Clin Infect Dis, 2025. 80(2): p. 389–396.

32. Conte, C., K.D. Hall, and S. Klein, Is Weight Loss-Induced Muscle Mass Loss Clinically Relevant? Jama, 2024. 332(1): p. 9–10.

33. Sanyal, A.J., et al., Triple hormone receptor agonist retatrutide for metabolic dysfunction-associated steatotic liver disease: a randomized phase 2a trial. Nat Med, 2024. 30(7): p. 2037–2048.

34. Golub, L.S., et al., The impact of semaglutide on liver fat assessed by serial cardiac CT scans in patients with type 2 diabetes: Results from STOP trial. Nutr Metab Cardiovasc Dis, 2025: p. 104036.

35. Gleason, P.P., et al., Real-world persistence and adherence to glucagon-like peptide-1 receptor agonists among obese commercially insured adults without diabetes. J Manag Care Spec Pharm, 2024. 30(8): p. 860–867.

36. Keys, A., et al., The biology of human starvation. (2 Vols). 1950: University of Minnesota Press.

37. Yates, T., et al., Impact of weight loss and weight gain trajectories on body composition in a population at high risk of type 2 diabetes: A prospective cohort analysis. Diabetes Obes Metab, 2024. 26(3): p. 1008–1015.

38. Osipov, B., et al., Sex differences in systemic bone and muscle loss following femur fracture in mice. Journal of Orthopaedic Research®, 2022. 40(4): p. 878–890.

39. Langer, H.T., et al., A mutation in desmin makes skeletal muscle less vulnerable to acute muscle damage after eccentric loading in rats. The FASEB Journal, 2021. 35(9): p. e21860.

